# Characterizing the transmission and identifying the control strategy for COVID-19 through epidemiological modeling

**DOI:** 10.1101/2020.02.24.20026773

**Authors:** Huijuan Zhou, Chengbin Xue, Guannan Gao, Lauren Lawless, Linglin Xie, Ke K. Zhang

## Abstract

The outbreak of the novel coronavirus disease, COVID-19, originating from Wuhan, China in early December, has infected more than 70,000 people in China and other countries and has caused more than 2,000 deaths. As the disease continues to spread, the biomedical society urgently began identifying effective approaches to prevent further outbreaks. Through rigorous epidemiological analysis, we characterized the fast transmission of COVID-19 with a basic reproductive number 5.6 and proved a sole zoonotic source to originate in Wuhan. No changes in transmission have been noted across generations. By evaluating different control strategies through predictive modeling and Monte carlo simulations, a comprehensive quarantine in hospitals and quarantine stations has been found to be the most effective approach. Government action to immediately enforce this quarantine is highly recommended.

## INTRODUCTION

The outbreak of the novel coronavirus disease, COVID-19, originated in Wuhan, a city located in central China, in early December, is fast spreading to more than 30 countries (Benvenuto et al., 2020; She et al., 2020). In less than 3 months, the disease has infected more than 70,000 people globally, and has caused more than 2,000 deaths. On January 30^th^, 2020, the World Health Organization declared COVID-19 as a public health emergency of international concern. Upon composing this manuscript, the coronavirus is concurrently spreading and claiming more than 100 lives per day. This outbreak induces an urgency to determine the characteristics of COVID-19 transmission and implement optimal strategies to control the epidemic.

Coronavirus is an enveloped, positive-sense, single stranded RNA virus found in a variety of mammals, including bats, civets, camel, and pangolins (Ge et al., 2013; Kandeil et al., 2019; Liu et al., 2007; Rockx et al., 2011). Two strains of coronaviruses, SARS-CoV-1 and MERS-CoV, have been reported to cause severe respiratory syndromes, resulting in deadly epidemics in 2002 and 2012, respectively (Fung and Liu, 2019; Luk et al., 2019). This current coronavirus, denoted as SARS-CoV-2, is the seventh known coronavirus to infect humans. (Benvenuto et al., 2020).

To effectively fight and end the COVID-19 epidemic, the transmission of the disease and zoonotic source of origin must be accurately identified, leading to the discovery of an optimal strategy to control this outbreak. By studying the cases and clinical features from early reports, this paper illustrates rigorous epidemiological models and the associated statistical methods to estimate the transmission rates in different stages and scenarios and predict the outcomes for different control strategies.

## METHODS

### Data sources

The numbers of cases from January 21^st^ to February 20^th^, 2020 were obtained from the daily reports by National Health Commission (NHC) of China. As NHC did not release case reports until January 21^st^, the case numbers prior to January 21^st^ were obtained from two recent clinical reports (Huang et al., 2020; Li et al., 2020). The number of patients diagnosed out of Wuhan was obtained from the report by (Guan et al., 2020) which summarized 1,099 cases confirmed by January 29^th^, 2020. The case numbers for the Diamond Princess Cruise were obtained from the daily reports by Yokohama Port Quarantine Center.

### SEIQ model

A susceptible-exposed-infectious-quarantine model was used for transmission analysis and prediction of epidemiological spread,

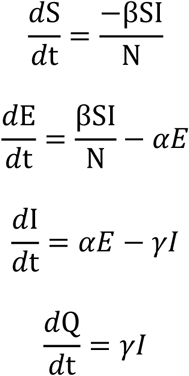

where S, E, I, Q and N were the number of susceptible (S), exposed (E), infectious (I), quarantined (Q) and total population. The population in Wuhan (N) is 11,081,000. Assuming that a patient was quarantined immediately after the diagnosis was confirmed, Q was equal to the confirmed number of cases. β is the daily transmission rate, defined as the expected number of infections caused by one infectious person per day. Once a susceptible (S) person becomes infected, the status is changed to exposed for an incubation period, 1/α. Theoretically, the patient is not infectious during the incubation period. After incubation, the patient experiences disease onset and becomes infectious (I). The time interval between disease onset to quarantine (Q) is the infectious time, 1/γ.

### Estimating transmission

Because the SEIQ model cannot be solved explicitly, β was estimated using the Monte carlo method, which simulated the two independent Poisson processes: daily exposed cases and the individual incubation time. To estimate β, there are two scenarios to consider. Firstly, the number of cases may only be available at the beginning and end of the study period. The examples used were the Diamond Princess Cruise and Wuhan prior to January 20^th^ when the diagnosis kit was not sufficiently available. In this scenario, β was estimated by Monte carlo approximation of the number of cases at the end of the targeted time period. Secondly, the number of Q is made available each day for the examples and the number of confirmed cases in China after January 20^th^. In this scenario, γ was first determined from clinical reports for the targeted time period, and then β was estimated by minimizing the mean squared errors from 1,000 Monte carlo samples that approximated the Q curve.

### Epidemics prediction

To evaluate different control strategies, Monte carlo samples were generated given β, γ, initial E, and initial I. The simulation was based on two independent Poisson processes: daily exposed cases and the individual incubation time. The daily means and 95% confidence intervals for Q were obtained over 1,000 Monte carlo runs.

## RESULTS

### The transmission in open cities

Epidemiological analysis was performed on data collected from the Mainland of China between January 1^st^ and January 20^th^, 2020. During this period, the suspected zoonotic source, the Huanan Seafood Market, was closed and travel restrictions were not yet enforced by the government. Based on the reported clinical analysis for the 425 cases prior to January 23^rd^, the average inoculation time interval was found to be 5.2 days and the average time interval from disease onset to a clinical visit was 5.8 days (Li et al 2020). In this study, the total number of cases, 8247, including both confirmed and suspected patients, on January 26, 2020 were used as the number of onsets (I+E) for January 20^th^, 2020. In the same way, the total number of cases reported between January 1^st^ and January 7^th^, which totaled 136, was used as I0 in the SEIQ model. Because the SEIQ model cannot be solved explicitly, Monte carlo simulation was exploited to determine the daily transmission rate β0=0.44 with 95% confidence interval (CI), 0.43 to 0.47 (Figure 1A). Given the mean time interval between onset and hospital quarantine of 12.5 days, the basic reproductive number, R0, was estimated to be 5.5 (95% CI 5.3 to 5.8).

**Figure 1.**
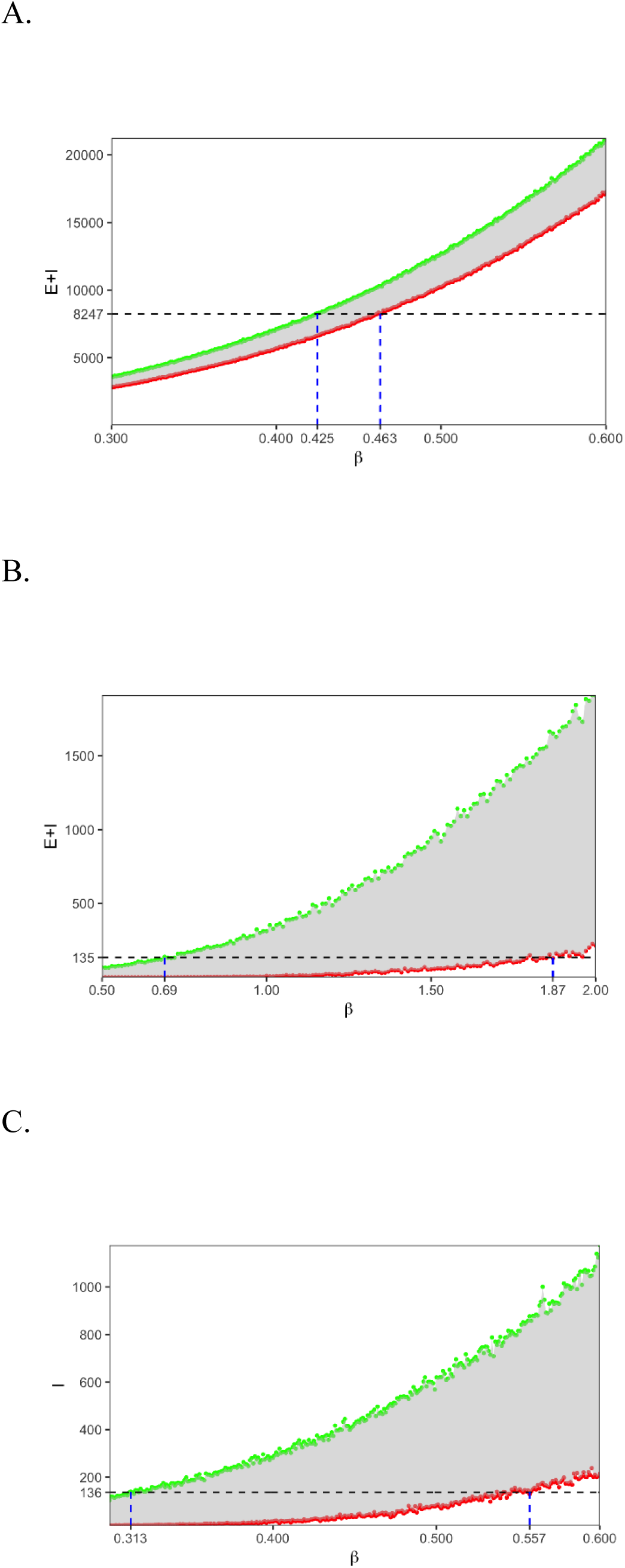
Estimation of the transmission rate β. For each β, 1,000 simulation runs were conducted and the low (green) and high (red) bounds of the 95% confidence intervals were plotted. The dashed blue lines indicated the 95% confidence interval of β for the targeted number of cases. **A**. Simulation of β for the time period from January 1^st^ to 20^th^ in China. 8247 was the total number of infectious (I) and exposed (E) on Jan. 20^th^. **B**. Simulation of β for the Diamond Princess Cruise. 135 was the total number of infectious (I) and exposed (E) on February 4^th^. **C**. Simulation of β for the time period from December 1^st^ to January 1^st^ in Wuhan. 135 was the number of infectious (I) on Jan. 1^st^.

### The transmission in an enclosed crowded environment

Most of infected cases in December 2019 were linked to the Huanan Seafood Market, which retails seafood and wild animals. It is believed that an enclosed and crowded environment is favorable for coronavirus transmission, however the epidemiological data is lacking. To test the hypothesis, the recent Diamond Princess cruise epidemic was used as a comparable case study. A clear disease outbreak was reported on this cruise. A passenger, who visited China on January 10^th^, was on board from January 20^th^ and January 25^th^ before being confirmed with a SARS-CoV-2 infection. All people on board have been quarantined at sea since February 5^th^ and 621 out of 3711 people were confirmed positive for SARS-CoV-2. As some of the patients were known to be infected after the quarantine, possibly due to central air conditioning and family infection, only the confirmed cases (n=135) by February 10^th^ (five days incubation plus one day diagnosis) were used for a conservative estimation of transmission. Using a SEIQ model while Q=0, the daily transmission rate, β, was found to be 1.04 (95% CI 0.69-1.87) (Figure 1B), which is twice as much as the transmission rate in open cities. Using the infection period of 12.5 days, the effective reproductive number on the cruise, Rc, was 13.0 (95% CI 8.63-23.375).

### A sole zoonotic source

The link between many of the December cases and the Huanan Seafood Market indicates that the Huanan Seafood Market is one of the zoonotic origins of SARS-CoV-2, if not only. After the forced shutdown of the Huanan Seafood Market on January 1^st^, the effectiveness of the zoonotic infection and a potential secondary source of SARS-CoV-2 that continued to infect the Wuhan people remained in question.

To characterize the zoonotic infection of COVID-19, a SEIQ model was constructed to analyze for the epidemic from the Hunan Seafood Market in December 2019. The first SARS-CoV-2 onset was found on December 1^st^ 2019 (Huang et al., 2020), and by the 1^st^ of January, 41 patients were confirmed and quarantined. Using the 5.8-day interval between disease onset and clinical visit, 136 cases diagnosed in the first 6 days of January were included, bringing the total number of infections to 177. Given β0=0.44, the development of the epidemic was simulated by initializing with a range of numbers (from 1 to 5) of infections on December 1^st^ using the SEIQ model. It showed that at most one infected patient can be allowed in the model, which induced a mean of 174 infections (95% CI 161-187) by January 1^st^. Instead of one, if two unrelated people were infected by December 1^st^, the 95% CI would be (291, 318) on January 1^st^, significantly larger than the expected 177 cases. Thus, the transmission would be so minimal that it would not substantially contribute to the final number of infections, even if there existed a second zoonotic source. In the same sense, the results did not support a continuous zoonotic source within the Huanan Seafood Market, which would have resulted in a higher overall number of infections in the later trajectory.

### No transmission variations between generations

As an RNA virus, coronavirus conveys a high mutation rate (Benvenuto et al., 2020), raising concerns whether the transmission would change between generations. The significantly higher mortality rate observed in Wuhan suggested that the transmission may attenuate over generations. To test this hypothesis, the patients infected in December 2019 were considered as the first generation because most of the cases had links to the Huanan Market, while the patients infected in January 2020 were considered the second-or-later generation. Given the data from December (assuming there is only one case at the beginning of December), β was estimated to be 0.41 (95% CI 0.31-0.56) (Figure 1C), which was not significantly different from the transmission rate β0=0.45, obtained between January 1^st^ and 20^th^. Furthermore, the transmission rates between Wuhan and other cities were also compared. Based on Guan *et al*’s analysis of 1,099 patients confirmed by January 29^th^ (Guan et al., 2020), 616 were identified outside of Wuhan. Out of the 616 patients193 had recently visited Wuhan. Using a mean infectious period of 5 days, β was calculated to 0.438 (β=(616-193)/193/5=0.438), which was within the 95% confidence interval of the previous estimation of β0 in Wuhan, 0.44 (95% CI 0.43-0.47). Therefore, no evidence of attenuation of transmission was found.

### Control strategies

Unprecedented measures were taken in Wuhan to stop the spread of COVID-19. Immediately after the official announcement of the novel coronavirus, home isolations and personal protection equipment such as face masks and gloves were enforced. Public transportation was limited and, eventually, all canceled. On January 23^rd^, the government suspended all plane, train and bus travel in and out of Wuhan. On February 2^nd^, a comprehensive quarantine strategy was taken by the Chinese government. All home-isolated patients were mandated to be hospitalized in the newly built square cabin hospitals, and all people who had suspected symptoms or had close contacts were demanded for mandatory isolation in the quarantine stations. The effects of these steps may not be clear until the end of the epidemic. Nevertheless, an intermediate analysis is needed for evaluating various control strategies.

Considering the one-day delay in the quarantine of the patients in the square cabin hospitals, the SEIQ model was used to fit the data into two phases, from January 23^rd^ to February 2^nd^, and from February 3^rd^ to February 20^th^. In the first phase, γ was chosen to be 1/6 based on the analysis by (Yang et al., 2020). Using least squared errors for an exhaustive search of β, the optimal transmission rate became 0.54, which was greater than the basic transmission rate β0=0.44 (Figure 2A). In the second phase, the time interval from disease onset to quarantine was assumed to drop by another 50% due to the implication of the square cabin hospitals. Thus, γ=1/6, the transmission rate β decreased to 0.10, which was an 81.5% reduction from the first phase and a 77.3% reduction from the basic transmission rate. In conclusion, the aggressive quarantine strategy of building square cabin hospitals has effectively decreased the transmission, whereas the usefulness of the travel ban, home isolation, and personal protection is still unclear.

**Figure 2.**
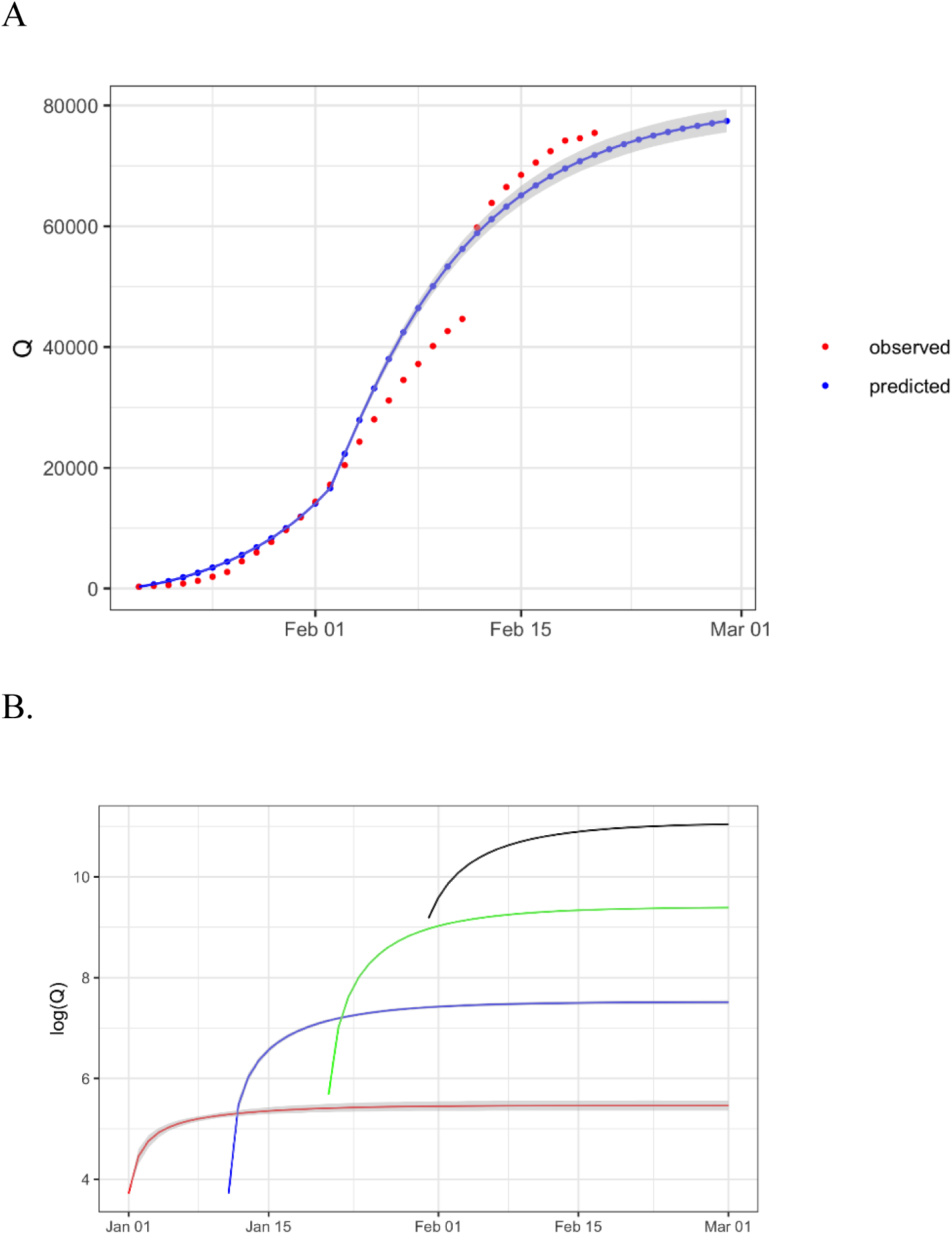
Simulation and prediction of the number of infections. **A**. The numbers of confirmed cases (Q) were predicted from the SEIQ model. The 95% confidence interval (gray area) were obtained from 1,000 Monte carlo runs. **B**. Simulations for quarantine were performed at 4 different starting dates: January 1^st^, 10^th^, 20^th^, and 30^th^. The predicted number of confirmed cases (Q) was plotted at the natural logarithm scale. The 95% confidence interval (gray area) for each curve was provided.

To further investigate the timing of the comprehensive quarantine, given β=0.10 and γ=1/6, simulated isolations were employed to track the numbers of confirmed cases (Q) at various starting dates, January 1^st^, 10^th^, 20^th^, and 30^th^. The initial number of cases was estimated by the SEIQ model. The predicted Q value was plotted on a logarithm scale up to February 29^th^ (Figure 2B). It was noted that all curves tended to stabilize after the quarantine measures, and the time interval to stabilization increased as the number of initial cases increased. Beginning the quarantine on January 1^st^, 136 initial cases were reported. Curve stabilization was achieved after about 10 days, with 235 cases reported at the end of this period. January 10^th^ started with approximately 1,250 cases and ended with 1,834 cases and January 20^th^ started with about 8,250 and ended with about 12,000. The January 30^th^ quarantine took almost 20 days to reach stabilization, and the final number of cases, 62,635, was nearly double the initial number of cases, 38,397. It was noted that the prediction was rather accurate as illustrated by the small 95% CI (gray area), and the CI decreased to negligible as the initial number of cases increased.

## DISCUSSION

COVID-19 is spreading at a much higher rate and at a larger scale compared to the 2003 SARS epidemic. The basic reproductive number, R0, for SARS was determined to be 3 with 95% CI being 2 to 4 (Dye and Gay, 2003). However, contrary to public knowledge, the earlier reports from the COVID-19 outbreak provided a R0 number less than or equal to that for SARS (Li et al., 2020; Wu et al., 2020; Zhou et al., 2020). This may have resulted from an inaccurate number of clinical cases due to delayed clinical visits, overloading of clinical resources, and a low sensitivity of the COVID-19 diagnosis kit. With more clinical reports published, the case numbers were adjusted based on the information presented, which gave a more accurate transmission estimation. The basic reproductive number for COVID-19 was then found to be 5.6, which is substantially higher than that for SARS.

In an enclosed and crowded environment, the transmission of SARS-CoV-2 was thought to significantly increase. The Diamond Princess Cruise provided an excellent case study as the development of COVID-19 on the cruise line has a clear infectious source, exposure time, quarantine time, and total number of infections. The transport of the virus through the central air conditioning system and the ineffective quarantine of the ship’s crew, however, raised some concerns. Nevertheless, a conservative estimation of transmission using only 50% of the confirmed cases has proven the high transmission rate in an enclosed crowded ship. This suggested that the initial offense within the Huanan Seafood Market could be at a high level.

The likely zoonotic origin also raised questions about additional sources other than the Huanan Seafood Market. This argument was partially supported by several early cases with no known links to the Huanan Seafood Market. Rigorous statistical models were used to test the hypothesis of a second zoonotic source, and the development of case numbers did not support this hypothesis. Though our model cannot completely exclude the possibility of additional zoonotic sources, it suggested a minimal effect by such sources. Furthermore, the model did not support the constant zoonotic infection within Huanan Seafood Market throughout December. If the zoonotic transgression was sustained, a larger number of infected cases would have been observed. Based on the data from the Diamond Process Cruise, the transmission rate was higher in an enclosed and crowded place, like the Huanan Seafood Market. However, similar transmission rate was observed in December as in January. This is likely due to the market’s daytime only operating hours, reducing the contact time. This result further suggested similar transmission rate between generations, which was confirmed by comparing the transmission in Wuhan and other cities. High mutation rates have been observed in the genome sequences collected from more than 100 human specimens. However, the effect that these mutations have on transmission may not be observed in such a short time period.

The Chinese government took unprecedented measures to fight the new epidemic and those measures raised global controversy for their necessity and effectiveness. The restrictive travel ban and home isolation enforced in Wuhan was expected to largely decrease the transmission and prevent further spread of the disease within weeks. Nevertheless, the coronavirus infection was continuously increasing exponentially in late January and early February in Wuhan. In this model, an increase in the transmission rate (from 0.44 to 0.54) was observed during this time period. It suggested that the travel ban and home isolation cannot effectively prevent the disease spread. This was likely due to the probable cross-contamination in the long waiting lines at the clinics and the contagion among family members. The limited medical resources only allowed patients with severe symptoms to be hospitalized. A low sensitivity of diagnosis further increased the waiting time for a confirmed case. Insufficient hospital beds resulted in a large number of home isolated patients, often leading to family infection. Observing the tremendous epidemic, the Chinese government built square cabin hospitals with more than twenty thousand beds and quickly moved all patients into these hospitals. All people with suspected symptoms or with close patient contacts were isolated in the government-managed quarantine stations. This comprehensive quarantine method has successfully reduced the transmission rate by 81.5%, and also greatly shortened the infectious time interval. The analysis in this study showed that the epidemic can be controlled within a few weeks if the comprehensive quarantine was conducted on January 20^th^ or earlier. Concurrently with the development of this manuscript, it was reported that South Korea had more than 1,000 home isolations in the city of Daegu for a suspected SARS-CoV-2 infection. It is highly recommended that other countries immediately quarantine all suspected patients.

This model was developed using the cases up to February 20^th^. As the case numbers increase, the transmission features may change and some of the assumptions, such as the infectious period, may vary. The cases reported in Wuhan were known to be less than the actual number in January due to overloading of clinical resources and the low sensitivity of diagnosis. This study attempted to overcome these limitations by using the clinical information verified by several reports and relying only on the beginning and ending cases. Therefore, this model tended to reflect the real trend of this critical epidemic and it provides more convincing evidence to guide the control of this disease by the government.

## Data Availability

All data used in this manuscript was obtained from public domain and published literature. The software code and simulated data are available upon request.

## REFERENCES

Benvenuto, D., Giovanetti, M., Ciccozzi, A., Spoto, S., Angeletti, S., and Ciccozzi, M. (2020). The 2019-new coronavirus epidemic: Evidence for virus evolution. J Med Virol 92, 455–459.

Dye, C., and Gay, N. (2003). Epidemiology. Modeling the SARS epidemic. Science 300, 1884-1885.

Fung, T.S., and Liu, D.X. (2019). Human Coronavirus: Host-Pathogen Interaction. Annu Rev Microbiol 73, 529–557.

Ge, X.Y., Li, J.L., Yang, X.L., Chmura, A.A., Zhu, G., Epstein, J.H., Mazet, J.K., Hu, B., Zhang, W., Peng, C., et al. (2013). Isolation and characterization of a bat SARS-like coronavirus that uses the ACE2 receptor. Nature 503, 535–538.

Guan, W.-j., Ni, Z.-y., Hu, Y., Liang, W.-h., Ou, C.-q., He, J.-x., Liu, L., Shan, H., Lei, C.-l., Hui, D.S., et al. (2020). Clinical characteristics of 2019 novel coronavirus infection in China. 2020.2002.2006.20020974.

Huang, C., Wang, Y., Li, X., Ren, L., Zhao, J., Hu, Y., Zhang, L., Fan, G., Xu, J., Gu, X., et al. (2020). Clinical features of patients infected with 2019 novel coronavirus in Wuhan, China. Lancet 395, 497–506.

Kandeil, A., Gomaa, M., Nageh, A., Shehata, M.M., Kayed, A.E., Sabir, J.S.M., Abiadh, A., Jrijer, J., Amr, Z., Said, M.A., et al. (2019). Middle East Respiratory Syndrome Coronavirus (MERS-CoV) in Dromedary Camels in Africa and Middle East. Viruses 11.

Li, Q., Guan, X., Wu, P., Wang, X., Zhou, L., Tong, Y., Ren, R., Leung, K.S.M., Lau, E.H.Y., Wong, J.Y., et al. (2020). Early Transmission Dynamics in Wuhan, China, of Novel Coronavirus-Infected Pneumonia. N Engl J Med.

Liu, L., Fang, Q., Deng, F., Wang, H., Yi, C.E., Ba, L., Yu, W., Lin, R.D., Li, T., Hu, Z., et al. (2007). Natural mutations in the receptor binding domain of spike glycoprotein determine the reactivity of cross-neutralization between palm civet coronavirus and severe acute respiratory syndrome coronavirus. J Virol 81, 4694–4700.

Luk, H.K.H., Li, X., Fung, J., Lau, S.K.P., and Woo, P.C.Y. (2019). Molecular epidemiology, evolution and phylogeny of SARS coronavirus. Infect Genet Evol 71, 21–30.

Rockx, B., Feldmann, F., Brining, D., Gardner, D., LaCasse, R., Kercher, L., Long, D., Rosenke, R., Virtaneva, K., Sturdevant, D.E., et al. (2011). Comparative pathogenesis of three human and zoonotic SARS-CoV strains in cynomolgus macaques. PLoS One 6, e18558.

She, J., Jiang, J., Ye, L., Hu, L., Bai, C., and Song, Y. (2020). 2019 novel coronavirus of pneumonia in Wuhan, China: emerging attack and management strategies. Clin Transl Med 9, 19.

Wu, J.T., Leung, K., and Leung, G.M. (2020). Nowcasting and forecasting the potential domestic and international spread of the 2019-nCoV outbreak originating in Wuhan, China: a modelling study. Lancet.

Yang, Y., Lu, Q., Liu, M., Wang, Y., Zhang, A., Jalali, N., Dean, N., Longini, I., Halloran, M.E., Xu, B., et al. (2020). Epidemiological and clinical features of the 2019 novel coronavirus outbreak in China. 2020.2002.2010.20021675.

Zhou, T., Liu, Q., Yang, Z., Liao, J., Yang, K., Bai, W., Lu, X., and Zhang, W. (2020). Preliminary prediction of the basic reproduction number of the Wuhan novel coronavirus 2019-nCoV. J Evid Based Med.

